# General Practice Perspectives on Post-Infection Conditions: Scoping Review and UK Survey

**DOI:** 10.64898/2026.07.15.26358157

**Authors:** Kalya Win Aung, Jamie Scuffell, Anna Podlasek, Serge Engamba, Fiona Jones, Adrian Edwards, Carolyn A. Chew-Graham, Leigh Sanyaolu, Monica Busse-Morris

## Abstract

**Background:** Post-infection conditions (PICs), such as Long Covid, are associated with heterogeneous, fluctuating symptoms that profoundly affect daily functioning. Despite moderate-certainty evidence from the NIHR-funded LISTEN trial (COV-LT2-0009) that personalised self-management support improves outcomes and may reduce societal and economic impacts of Long Covid, many people living with PICs still receive condition-specific services, generic advice, or stand-alone digital tools that do not address their complex needs.

**Aim:** To map care approaches in general practice and synthesise UK evidence for PIC management.

**Design and setting:** Scoping review and online survey.

**Method:** A two-phase study was conducted: (1) a scoping review of UK evidence on PIC management in general practice; and (2) a supplementary online survey of practitioners working in UK general practice to provide contextual insights.

**Results:** The scoping review identified 32 studies focused on Long Covid. One study included a comparator group (ME/CFS). Study populations were predominantly white ethnicity and female. Evidence for non-Covid PICs in UK general practice was largely absent.

The supplementary survey (n=46) provided preliminary practice-level insights. Healthcare practitioners reported varied PIC presentations, diagnostic uncertainty, limited referral pathways, inequitable access, and low confidence in managing PICs.

**Conclusion:** Evidence informing PIC management in UK general practice remains predominantly Long Covid-focused and may not reflect the range of PICs encountered in practice. While survey findings are preliminary and require confirmation in larger samples, they highlight uncertainty around PIC management. Further research is needed to evaluate whether existing Long Covid pathways should be expanded or complemented by broader PIC models.

**How this fits in:** People living with PICs often seek support in general practice, yet the evidence base informing management remains predominantly focused on Long Covid, with limited evidence for other PICs. This study provides preliminary insights into potential challenges in applying current approaches across the wider PIC landscape. While self-management support was a common feature of reported care models, approaches were variable and often focused on generic advice or stand-alone digital resources rather than personalised, relational support. Further research is needed to understand how self-management support can be better delivered to help people feel believed and supported to navigate living with a PIC.

## Introduction

Acute infections are considered short-term illnesses, with most individuals expected to make a full recovery.^1^ However, a proportion experience persistent symptoms sometimes lasting months or longer. In the context of Covid-19, a recent global meta-analysis estimated that 36% of individuals report ongoing symptoms consistent with Long Covid.^2^ Similar longer-term symptoms have also been reported following other infections, including Lyme disease,^3^ dengue,^4^ and pneumonia.^5^ In this paper, we refer to these longer-term effects as post-infection conditions (PICs).

General practice is the first point of contact for most people living with PICs and is central to ongoing management.^6^ However, PIC presentations are heterogeneous and therefore difficult to classify within existing disease frameworks.^7^ Although PICs have long been recognised in general practice, approaches to identification and management are reported to be inconsistent.^8^ This lack of standardisation is compounded by limited evidence-based treatment options and the absence of structured, cross-condition pathways.^1,9^

Self-management is widely recognised as a key component of care for long-term conditions and is particularly relevant in PICs, where individuals must manage fluctuating symptoms over time.^10,11^ Evidence from the NIHR-funded LISTEN programme demonstrated that personalised self-management support can improve outcomes for people living with Long Covid and deliver wider societal and economic benefits.^12^ However, many people still report receiving generic advice or fragmented signposting rather than structured, personalised support embedded within routine care.^13–15^

While the evidence base for Long Covid is expanding, there is limited knowledge about how personalised self-management support is delivered across broader PICs in UK general practice. This study combines a scoping review with a small sample of clinician perspectives to contextualise current practice.

Within the scope of people living with PICs, the objectives were to:

- describe how self-management support is delivered;
- explore clinician perspectives on current practice;
- produce preliminary summaries of evidence on coding and management in UK general practice.

## Methods

A mixed-methods scoping study was undertaken in two phases. Ethical approval was obtained from the King’s College London College Research Ethics Committee (MRA-25/26-54995).

### Phase 1: Scoping review

#### Design

A scoping review was conducted in accordance with the Joanna Briggs Institute (JBI) methodology for scoping reviews^16^ and reported following PRISMA-ScR guidance.^17^ The review question and eligibility criteria were structured using the Population-Concept-Context (PCC) framework (Table 1).

**Table 1.** PCC framework for developing the research question and search strategy.

|  |  |
| --- | --- |
| Population | Adults (≥18 years) with PICs, including Long Covid, post-viral fatigue syndromes, ME/CFS with an infectious trigger, and related post-infectious sequelae |
| Concept | Primary care/general practice management, self-management support, care models, and experiences of care |
| Context | UK primary care, general practice, primary care networks, and community health services |

#### Search strategy

An initial limited search of MEDLINE and CINAHL was conducted to identify relevant articles and refine search terms. A comprehensive search was then conducted on 6 March 2026 across MEDLINE, Embase, APA PsycINFO, CINAHL, and the Cochrane Library. Grey literature was searched using The King’s Fund Library database, and reference lists of included studies and relevant reviews were screened for further eligible studies. Searches were limited to English-language publications. No date limits were applied. The full search strategy can be found in Supplementary Table 1.

#### Study selection

Records were imported into Covidence (Veritas Health Innovation, Melbourne, Australia) for deduplication. Titles, abstracts, and full texts were screened independently by two reviewers (KWA and MBM), with disagreements resolved through discussion. Consensus AI (https://consensus.app/) was used as a supplementary tool to support identification of missing studies.

#### Data extraction

Data were extracted independently by two reviewers (KWA and MBM) using a structured, piloted extraction form developed for this review. No formal critical appraisal was undertaken.

### Phase 2: National survey

#### Design

A cross-sectional online survey was conducted concurrently with the scoping review to explore the perspectives of healthcare practitioners working in UK general practice on PIC management, including self-management support and care delivery approaches. Findings were used alongside the scoping review to provide complementary contextual insight and to triangulate emerging evidence.

#### Survey development

Survey content was informed by existing literature on Long Covid and post-viral conditions^8,18,19^ and refined following consultation within the multidisciplinary research team. The survey was designed using Qualtrics (XM) software (Provo, UT), and included multiple-choice, Likert-scale, checkbox, and optional free-text questions.

Adaptive branching logic was used to tailor questions by professional role: general practitioners (GPs) received items focused on diagnosis, investigation, and coding, while other primary care practitioners were asked about management approaches, rehabilitation, and self-management support. Both versions included questions relating to service provision, referral pathways, confidence, and future workforce models. The survey was piloted with the research team and practitioners from the Primary Care Academic CollaboraTive (PACT) network, with minor refinements made prior to dissemination. Full versions of both surveys are provided in Supplementary Box 2.

#### Participants and recruitment

Eligible participants were UK-based practitioners working in general practice who were involved in the assessment and/or management of people with post-infectious symptoms, including GPs, advanced nurse practitioners, nurses, physiotherapists, pharmacists, and physician associates. Recruitment was conducted via national professional and academic networks, such as the PACT network (https://www.gppact.org/) and the TARGET (Treat Antibiotics Responsibly, Guidance, Education, and Tools) mailing list (https://www.rcgp.org.uk/TARGETantibiotics), hosted by the UK Health Security Agency (UKHSA). Participation was voluntary.

#### Data collection

The survey was open from March to May 2026 and took approximately 10-15 minutes to complete. Participants provided electronic informed consent via a secure online survey link. Responses were anonymised or pseudonymised depending on recruitment route.

Participants were offered entry into a £100 shopping voucher prize draw as an incentive for participation. Participants recruited through the PACT network also received an individualised practice feedback report.

#### Data analysis

Quantitative data were analysed descriptively and stratified by practitioner type (GP and other primary care health practitioners). Responses were summarised using frequencies, proportions, and descriptive statistics.

Qualitative free-text responses were analysed using inductive thematic analysis.^20^ Two authors (KWA and MBM) independently coded responses, developed preliminary codes, and refined themes through discussion. Analysis was iterative and informed by the study aims, with disagreements resolved by consensus.

#### Estimation of GP workload associated with PICs

Survey responses were also used to inform an exploratory estimate of coding related to PICs in general practice in England. In UK general practice, SNOMED-CT codes are used to record clinical activity and diagnoses.

A SNOMED-CT code list was derived for each of the responses to the question: “Upon seeing a person with a post-infection condition, which of these codes may you use to code the consultation?” This was created with a semantic and keyword search of the UK SNOMED-CT database. Codes were manually reviewed by a GP familiar with general practice in England (JS). The resulting code list was linked to NHS England data on coding in English primary care from 2011 to 2025, made available through the OpenCodeCounts R package.^21^

Analysis was performed in R (v4.4.1), with some coding support from Claude Opus 4.8 (Anthropic, San Francisco, USA). All code and outputs were checked by the authors.

## Results

### Phase 1: Scoping review

#### Search process

The study selection process is shown in Figure 1. Database searches yielded 430 records. After removal of duplicates, 295 records were screened, of which 233 were excluded at title/abstract stage. Sixty-two full-text articles were assessed for eligibility, with 34 excluded due to not meeting inclusion criteria. Four additional eligible studies were identified through Consensus AI. In total, 32 studies were included in the final synthesis.

**Figure 1.**
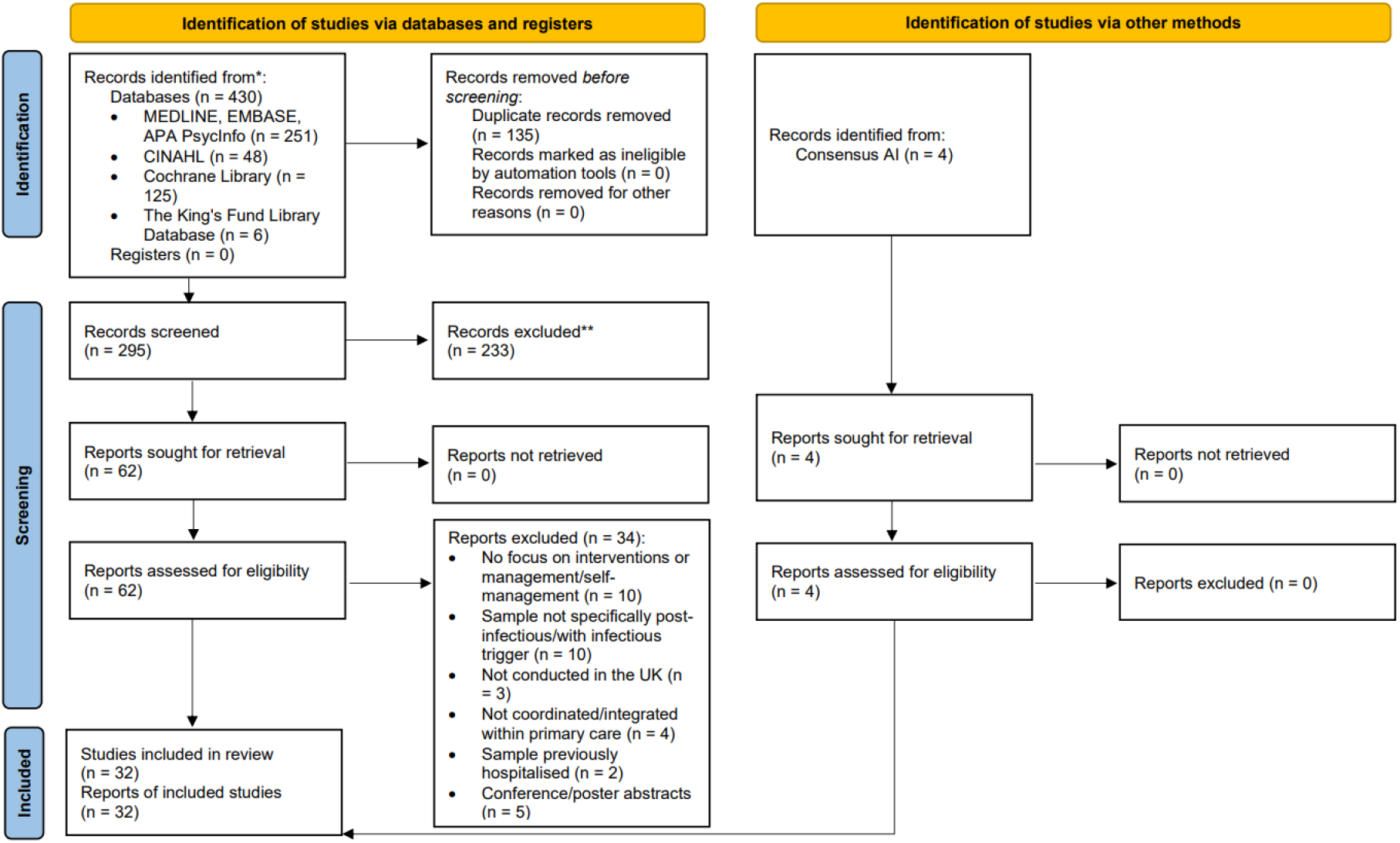
PRISMA flow diagram of literature search and screening results.

#### Paper characteristics

All 32 studies were published from 2020 onwards and focused exclusively on Long Covid, with no eligible evidence identified for other PICs such as post-Lyme disease, Epstein-Barr virus-related syndromes, or non-Covid post-viral fatigue. One study included a comparator group of people with ME/CFS alongside a Long Covid group but there was no confirmed infectious trigger at individual participant level.^22^ Study designs were predominantly mixed-methods (25%), qualitative (22%), and randomised controlled trials (RCTs) or RCT protocols (22%). Study settings included primary care, NHS Long Covid clinics, community services, and digital or hybrid models. Data extraction is presented in full in Supplementary Table 3.

#### Narrative synthesis

##### Care models and general practice coordination

GP-led care was consistently positioned as the entry point for Long Covid management across national guidance and expert consensus statements.^1,10,23^ These studies describe GPs as responsible for initial assessment, exclusion of alternative diagnoses, and coordination of referral to specialist services and digital self-management platforms.

##### Multidisciplinary and integrated care

A smaller number of studies describe multidisciplinary team (MDT) models involving physiotherapists, psychologists, nurses, and occupational therapists. These models generally report improved coordination and more holistic care.^23–26^ However, they are mainly focused on Long Covid and vary in implementation, suggesting that the extent to which these models are integrated into care is inconsistent.

##### Self-management support and digital delivery

Self-management support is consistently described as a key part of care across studies. This usually includes support for strategies such as pacing, symptom monitoring, and behavioural or rehabilitation-based approaches delivered through structured programmes or digital platforms such as NHS “Your COVID Recovery”.^10,12,27^ While these approaches are perceived to be generally acceptable, several studies highlight that digital delivery may not be accessible to all, particularly those with lower digital literacy or limited resources.^28,29^

##### Patient experience and equity

Across qualitative and survey studies, individuals commonly report fragmented care, difficulty getting a diagnosis, and feeling that symptoms are not always taken seriously.^30,31^ An important group of studies highlights inequities in access to care among minority ethnic and socioeconomically disadvantaged groups.^28,32^

##### Facilitators and barriers

Barriers such as service fragmentation, limited training, and workforce constraints were reported across qualitative studies^29,33,34^ and service evaluations.^24^ Facilitators including MDT working, social prescribing, and structured digital tools were identified in intervention and policy studies.^1,10,24,25^ Overall, the evidence suggests that care models are emerging, but their implementation remains inconsistent.

### Phase 2: National survey

#### Respondent characteristics

A total of 46 healthcare practitioners completed the survey, including 34 GPs (74%). Respondents were predominantly based in England (89%), with broad distribution across multiple Integrated Care Boards and no dominant regional clustering. Full demographic and practice characteristics are shown in Table 2.

**Table 2.** Demographic and practice characteristics of survey respondents.

| **Domain** | **Category** | **n (%)** |
| --- | --- | --- |
| Total sample | All respondents | 46 (100) |
| Role | GP | 34 (74) |
|  | Other healthcare practitioners | 11 (24) |
|  | Unspecified | 1 (2) |
| GP role type | GP Partner | 14 (41)* |
|  | Salaried GP | 14 (41)* |
|  | GP Registrar | 3 (9)* |
|  | Locum GP | 3 (9)* |
| Working pattern | >7 sessions/week | 2 (4) |
|  | 5-7 sessions/week | 21 (46) |
|  | 2-4 sessions/week | 22 (48) |
|  | <2 sessions/week | 1 (2) |
| Geographical distribution | England | 41 (89) |
|  | Scotland | 2 (4) |
|  | Wales | 3 (7) |
|  | Northern Ireland | 1 (2) |
|  | Unresolved postcodes | 5 (11) |

\*Percentages within GP sample (n=34).

#### Exposure to PICs in practice

PICs were commonly encountered in routine practice, with most GPs (85%) reporting at least one case in the preceding month (Table 3).

**Table 3.**
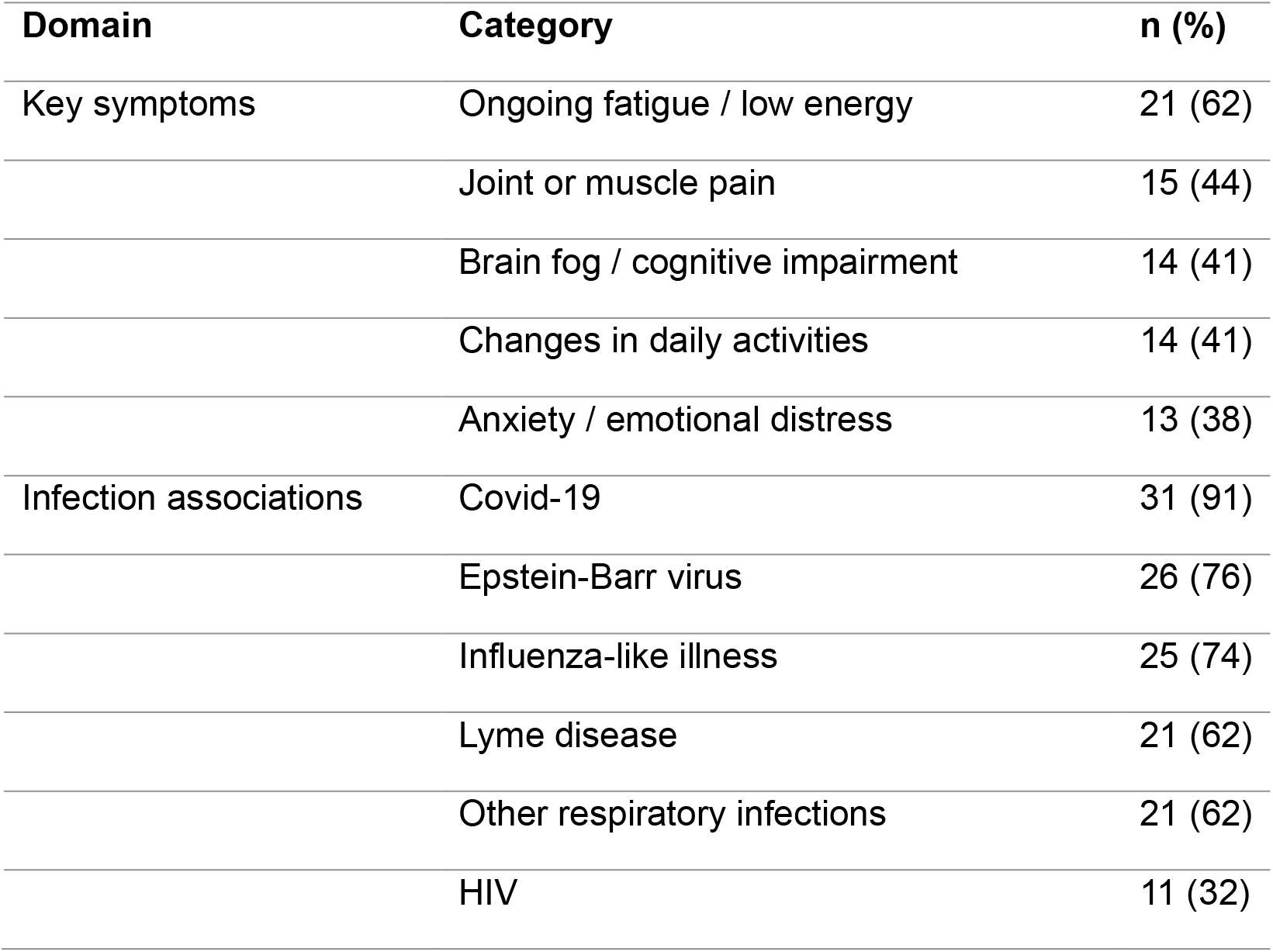
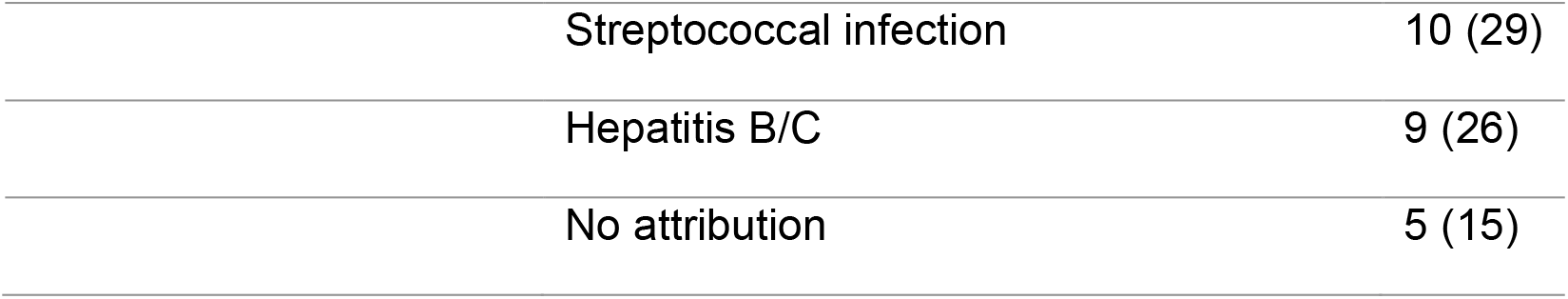
GP-reported symptom patterns and infections associated with PICs.

| Domain | Category | n (%) |
| --- | --- | --- |
| Key symptoms | Ongoing fatigue / low energy | 21 (62) |
|  | Joint or muscle pain | 15 (44) |
|  | Brain fog / cognitive impairment | 14 (41) |
|  | Changes in daily activities | 14 (41) |
|  | Anxiety / emotional distress | 13 (38) |
| Infection associations | Covid-19 | 31 (91) |
|  | Epstein-Barr virus | 26 (76) |
|  | Influenza-like illness | 25 (74) |
|  | Lyme disease | 21 (62) |
|  | Other respiratory infections | 21 (62) |
|  | HIV | 11 (32) |

|  |  |
| --- | --- |
| Streptococcal infection | 10 (29) |
| Hepatitis B/C | 9 (26) |
| No attribution | 5 (15) |

#### Common symptoms of PICs and associated infections

GPs consistently described PICs as multi-system in nature with fluctuating presentations. Symptoms reported “about half the time” or more commonly included fatigue, musculoskeletal pain, and cognitive difficulties. Covid-19 was the infection most commonly associated with persistent symptoms (Table 3).

#### Clinical assessment and management confidence

When people presented with these symptoms at initial consultations, most GPs reported routine history-taking and physical examination at first presentation. Follow-up consultations placed greater emphasis on targeted investigations and rehabilitation referrals at follow-up (Table 4).

**Table 4.**
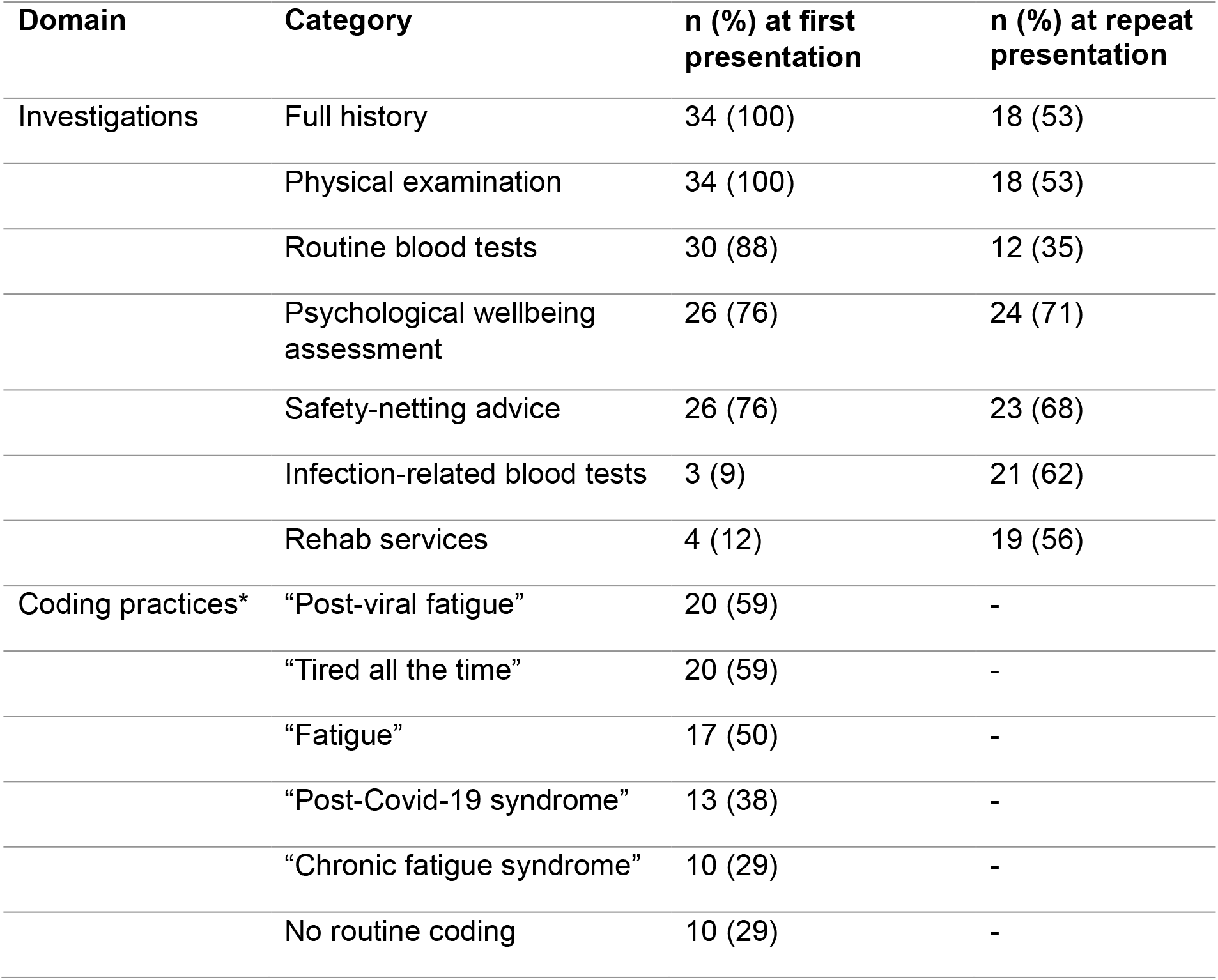
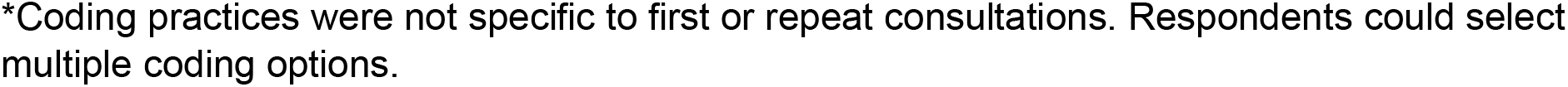
GP-reported clinical investigations and coding practices for PICs.

| **Domain** | **Category** | **n (%) at first presentation** | **n (%) at repeat presentation** |
| --- | --- | --- | --- |
| Investigations | Full history | 34 (100) | 18 (53) |
|  | Physical examination | 34 (100) | 18 (53) |
|  | Routine blood tests | 30 (88) | 12 (35) |
|  | Psychological wellbeing assessment | 26 (76) | 24 (71) |
|  | Safety-netting advice | 26 (76) | 23 (68) |
|  | Infection-related blood tests | 3 (9) | 21 (62) |
|  | Rehab services | 4 (12) | 19 (56) |
| Coding practices* | “Post-viral fatigue” | 20 (59) | - |
|  | “Tired all the time” | 20 (59) | - |
|  | “Fatigue” | 17 (50) | - |
|  | “Post-Covid-19 syndrome” | 13 (38) | - |
|  | “Chronic fatigue syndrome” | 10 (29) | - |
|  | No routine coding | 10 (29) | - |

\*Coding practices were not specific to first or repeat consultations. Respondents could select multiple coding options.

When considering a PIC, most GPs (85%) said they did not apply a fixed diagnostic timeframe, instead relying on symptom duration and clinical judgement. Confidence in managing PICs was mixed, with 50% of GPs and 55% of other healthcare practitioners reporting low confidence in their ability to manage these presentations.

#### Coding patterns and exploratory workload estimation

Coding practices were inconsistent, with 29% of GPs reporting that they did not routinely code consultations for PICs. Where coding was used, respondents reported multiple overlapping symptom-based and diagnostic labels, reflecting heterogeneous approaches to recording PIC-related consultations (Table 4).

Exploratory analysis of SNOMED-CT coding data indicated that symptom-based codes such as fatigue, chronic pain, and “tired all the time” accounted for the majority of recorded activity in general practice in England, while codes explicitly naming PICs were used less frequently and showed temporal variation. Full distributions are presented in Supplementary Figure 4.

#### Service access and perceived gaps

Referral pathways were inconsistent, with 44% of GPs reporting structured pathways and 38% reporting none. Services were most commonly mental health (80%) and community rehabilitation (67%), while access to dedicated PIC services was rare. Most GPs (82%) felt current services met needs only to a limited extent or not at all.

There was support from general practice practitioners for more personalised, multidisciplinary models of care, with most GPs (88%) and other healthcare practitioners working in general practice (91%) reporting that such services would be beneficial to at least some extent.

#### Thematic analysis of free-text responses

Free-text responses identified five interlinked themes relating to a lack of clear pathways for PICs, diagnostic ambiguity, limited treatment options, reliance on self-management, and the need for more integrated and personalised care (Table 5).

**Table 5.**
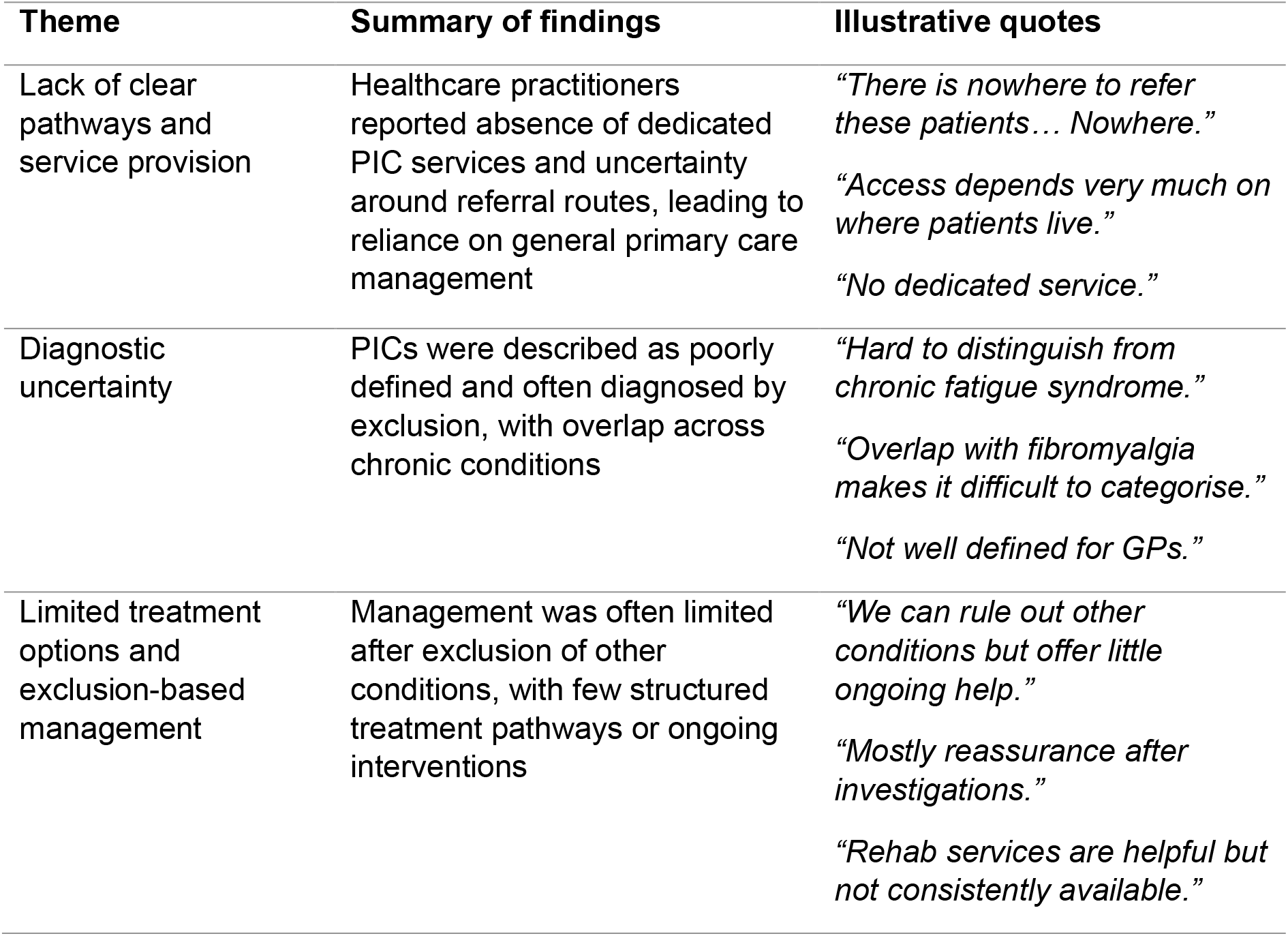
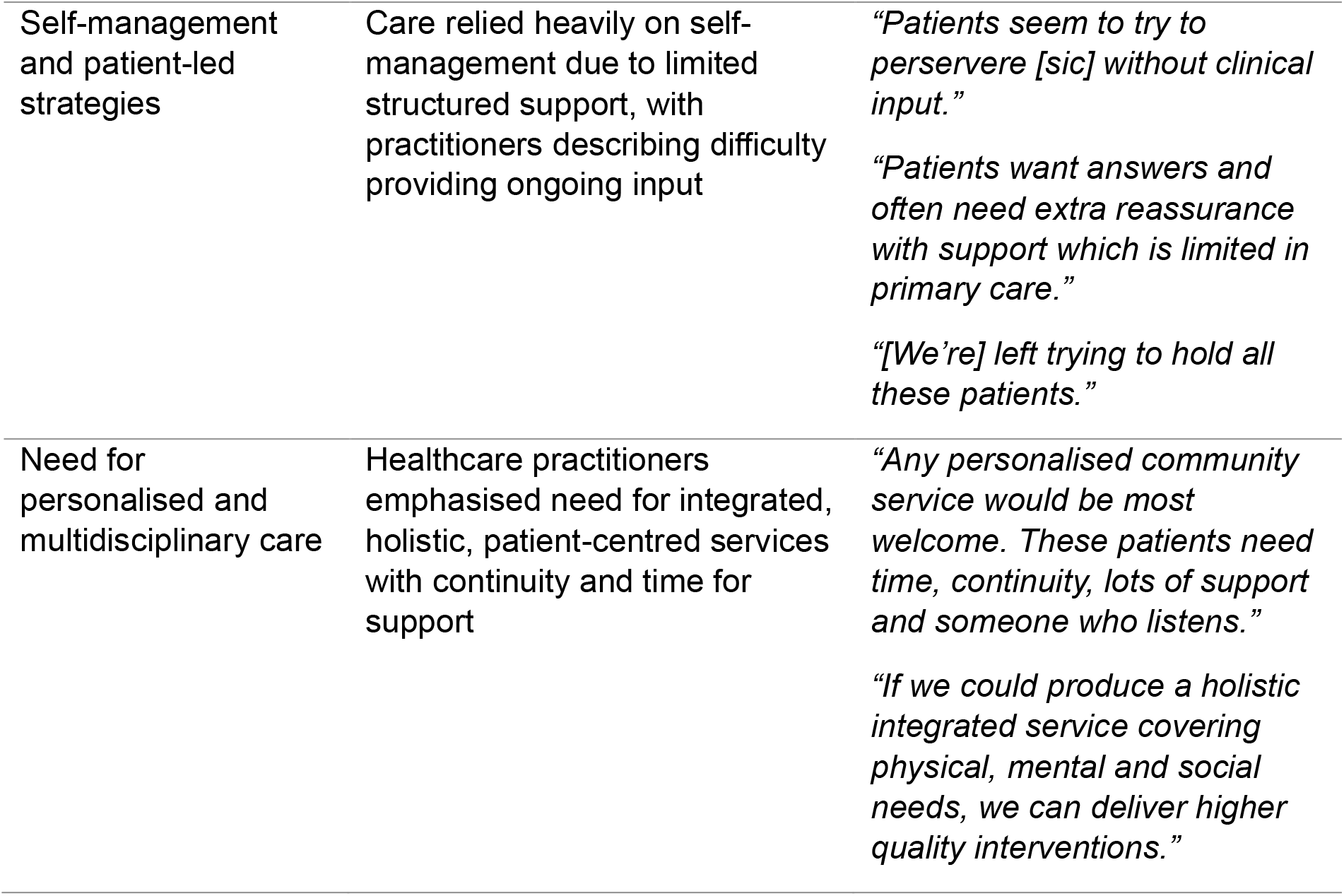
Thematic analysis of healthcare practitioners’ free-text responses on PIC management.

## Discussion

### Summary

This study combined a scoping review and preliminary survey to examine care delivery and self-management support for PICs in the UK. The scoping review (n=32) showed that the evidence-base is almost entirely focused on Long Covid, with no eligible studies addressing other PICs beyond a single ME/CFS comparator group.^22^ Across studies, general practice was central to assessment and coordination, while integrated multidisciplinary and digital self-management approaches were commonly proposed. However, implementation was variable, and evidence on equitable access and outcomes remained limited despite concerns about disparities in service access and digital exclusion.

Survey findings (n=46) provided complementary contextual insight, suggesting healthcare practitioner uncertainty and low confidence in the management of PICs in general practice. Consistent with the scoping review, healthcare practitioners reported fragmented service provision, workforce constraints, and limited access to integrated multidisciplinary care. Despite this, they consistently emphasised the importance of multidisciplinary and personalised approaches, with self-management seen as central but often insufficiently supported in routine general practice.

### Strengths and limitations

This study integrates two complementary evidence sources, strengthening understanding of both real-world clinical practice and the broader research landscape. The mixed-methods design allowed triangulation between clinician experience and published evidence, which indicated convergence of findings. However, the survey sample was small (n=46) and not representative of UK primary care. Despite this, linkage to routine data sources allowed exploratory estimates of England-wide PIC-related coding activity, although these are based on codes recorded for any indication rather than patient-level PIC diagnoses and therefore likely overestimate true PIC-specific activity. The scoping review was restricted to English-language studies and was dominated by Long Covid research, limiting generalisability to other PICs. Heterogeneity in study designs and outcomes also limited comparability.

### Comparison with existing literature

Prior literature has largely focused on Long Covid as the dominant PIC within policy and research, with general practice framed as the entry point for coordination of assessment and referral pathways.^1,6,7,10,11^ Our scoping review aligns with this pattern, identifying no eligible UK general practice studies addressing other PICs. However, our survey findings, while limited by sample size, indicated that general practitioners and other health professionals working in general practice encounter a wider range of post-infectious presentations in routine practice. This highlights a potential gap between the current research focus and clinical need, although further research with more representative samples is required to confirm this.

### Implications for research and/or practice

The evidence base and service models for PIC management are largely shaped by Long Covid, while approaches for other PICs are less developed. As dedicated Long Covid services diminish, it remains unclear whether future care should adapt existing Long Covid pathways or develop broader alternative service models. Research should therefore prioritise comparative evaluation of different approaches to service organisation, including the role of structured self-management support, with attention to effectiveness, equity, workforce confidence, and continuity of care across PIC populations.

## Data Availability

All data produced in the present study are available upon reasonable request to the authors.

## Ethical approval

Ethical approval was obtained from the King’s College London College Research Ethics Committee (MRA-25/26-54995).

## Acknowledgements

We gratefully acknowledge the healthcare professionals who generously shared their time, experiences, and perspectives, as well as the organisations and networks that supported recruitment for this study. We particularly thank the following survey respondents who consented to be acknowledged by name: Hannah Ferrington, Antony Willman, Imran Rauf, Adnan Ezouli, Anna Podlasek, Nes Robson, Mark Thomas, Alaa Awad, Dr Laura Smith, Dr Olajide Popoola, Loukas Kouzaris, Dr I Ghafoor, Alistair Jones, Deepthi Lavu, Rob Audain, Dr Emma Tonner, Victoria Hodges, Laura Parrish, Deian Howarth, Rachel Wallbank, Kelly Quirk, Dr Rania El-Beltagy, Dr Kieran McCormack, and Emma Horner. We are equally grateful to all other healthcare professionals who participated in the survey and contributed to this research.

