## Supplementary Materials S1-4 for "General Practice Perspectives on Post-Infection Conditions: Scoping Review and UK Survey"

**Supplementary Data**

**Supplementary Table S1. Final search strategies for electronic databases and grey literature sources.**

| Database / source | Date searched | Limits / restrictions | Search results | Search strategy |
| --- | --- | --- | --- | --- |
| MEDLINE, EMBASE, APA PsycInfo (via Ovid) | 6 March 2026 | No limits / restrictions | 251 | Concept 1: Post-infection conditions (long-covid* or post-covid* or PASC or myalgic encephalomyelitis or chronic fatigue syndrome or "ME/CFS" or CFS or post-viral or post viral or post-viral syndrome* or post-infect* or post infectious syndrome* or post-acute infect* or post-acute covid*) AND Concept 2: General practice / primary care (primary care or primary health care or primary healthcare or general practitioner* or general practic* or GP or general medical practice or family practi* or family physician or family practitioner or family medicine or primary care network* or PCN or community health cent*) AND Concept 3: Management and support approaches (self-management or self-manag* or self-care or self-regulation or self-monitoring or health coaching or social prescrib* or personalised care or personalized care or care coordination or care model or service model or service delivery or rehabilitation or treatment or intervention*) AND Concept 4: UK setting (United Kingdom or UK or England or Scotland or Wales or Northern Ireland or NHS) |
| CINAHL (via EBSCOhost) | 6 March 2026 | No limits / restrictions | 48 | (long-covid* or post-covid* or PASC or myalgic encephalomyelitis or chronic fatigue syndrome or "ME/CFS" or CFS or post-viral or post viral or post-viral syndrome* or post-infect* or post infectious syndrome* or post-acute infect* or post-acute covid*) AND (primary care or primary health care or primary healthcare or general practitioner* or general practic* or GP or general medical practice or family practi* or family physician or family practitioner or family medicine or primary care network* or PCN or community health cent*) AND (self-management or self-manag* or self-care or self-regulation or self-monitoring or health coaching or social prescrib* or personalised care or personalized care or care coordination or care model or service model or service delivery or rehabilitation or treatment or intervention*) AND (United Kingdom or UK or England or Scotland or Wales or Northern Ireland or NHS) |
| Cochrane Library | 6 March 2026 | No limits / restrictions | 125 | (long-covid* or post-covid* or PASC or myalgic encephalomyelitis or chronic fatigue syndrome or "ME/CFS" or CFS or post-viral or post viral or post-viral syndrome* or post-infect* or post infectious syndrome* or post-acute infect* or post-acute covid*) AND (primary care or primary health care or primary healthcare or general practitioner* or general practic* or GP or general medical practice or family practi* or family physician or family practitioner or family medicine or primary care network* or PCN or community health cent*) AND (self-management or self-manag* or self-care or self-regulation or self-monitoring or health coaching or social prescrib* or personalised care or personalized care or care coordination or care model or service model or service delivery or rehabilitation or treatment or intervention*) AND (United Kingdom or UK or England or Scotland or Wales or Northern Ireland or NHS) |
| The King's Fund Library Database | 6 March 2026 | No limits / restrictions | 6 | ("long covid" OR "post covid" OR "post viral fatigue" OR "chronic fatigue syndrome" OR "myalgic encephalomyelitis" OR "post viral" OR "post infection" OR "post infectious" OR "following infection" OR "after infection") AND ("primary care" OR "primary health care" OR "general practice" OR GP OR "general practitioner" OR "general medical practice" OR "family practice" OR "family practitioner" OR "family medicine" OR "primary care network" OR PCN OR "community health centre" OR "community health center") AND ("self management" OR "self care" OR "self regulation" OR "self monitor" OR "self monitoring" OR "health coaching" OR "social prescribing" OR "personalised care" OR "personalized care" OR "care coordination" OR "care model" OR "service model" OR "service delivery" OR rehabilitation OR treatment OR intervention) AND ("United Kingdom" OR UK OR England OR Scotland OR Wales OR "Northern Ireland" OR NHS) |

**Supplementary Box S2. Full GP and other healthcare practitioner survey.**

Survey 1: General Practitioner (GP) Survey

Practice name: _______________________

Practice postcode: _______________________

Role

☐ GP Partner

☐ Salaried GP

☐ Locum GP

☐ GP Registrar

Number of patient-facing sessions per week

☐ <2 sessions

☐ 2-4 sessions

☐ 5-7 sessions

☐ 8 sessions

Main survey

This survey includes 15 questions and takes approximately 10-15 minutes to complete.

Definition: Post-infection conditions (PICs) describe persistent, episodic, or recurrent multi-system symptoms following an acute infection. PICs include, but are not limited to, Long Covid.

1. In the last month, how many patients have you seen with persistent, episodic, or recurrent multi-system symptoms that you and/or the patient attributed to a prior infection?

☐ None
☐ 1–5
☐ 6–10
☐ 11–20
☐ >20
☐ Not sure

2. How often would you attribute the following symptoms to a post-infection condition in your patients?

| Symptom | Always | Most of the time | About half the time | Sometimes | Never |
| --- | --- | --- | --- | --- | --- |
| Ongoing fatigue and low energy | ☐ | ☐ | ☐ | ☐ | ☐ |
| Brain fog, memory problems, or difficulty concentrating | ☐ | ☐ | ☐ | ☐ | ☐ |
| Chest pain on exertion | ☐ | ☐ | ☐ | ☐ | ☐ |
| Chest pain at rest | ☐ | ☐ | ☐ | ☐ | ☐ |
| Anxiety or emotional distress | ☐ | ☐ | ☐ | ☐ | ☐ |
| Headaches or migraines | ☐ | ☐ | ☐ | ☐ | ☐ |
| Joint or muscle pain | ☐ | ☐ | ☐ | ☐ | ☐ |
| Changes in mobility | ☐ | ☐ | ☐ | ☐ | ☐ |
| Changes in ability to complete usual activities | ☐ | ☐ | ☐ | ☐ | ☐ |
| Changes in appetite or digestive function | ☐ | ☐ | ☐ | ☐ | ☐ |
| Palpitations, dizziness, or light-headedness on standing | ☐ | ☐ | ☐ | ☐ | ☐ |

3. Would you attribute any other symptoms not listed above to a PIC?

☐ Yes
☐ No

If yes, please specify: _______________________

4. What factors might make you more likely to attribute symptoms to a PIC? (Select all that apply)

☐ Women
☐ Older adults
☐ Smoking
☐ Overweight/obesity
☐ Pre-existing chronic health conditions
☐ Disability
☐ Repeated infections
☐ Previous hospitalisation/ICU admission
☐ Other: __________
☐ None of the above

5. Which infections do you associate with persistent, episodic, or recurrent multi-system symptoms?

☐ COVID-19
☐ Influenza/flu-like illness
☐ Epstein–Barr virus (glandular fever)
☐ Lyme disease
☐ Respiratory infections (e.g., pneumonia, pertussis)
☐ Other bacterial infections
☐ Streptococcal infections
☐ Hepatitis B/C
☐ HIV
☐ Cytomegalovirus
☐ I do not routinely attribute symptoms to a specific infection
☐ Other: __________

6-7. When adults present with the following symptoms, which actions are you most likely to take?

(Asked separately for first presentation and repeat attendance with ongoing symptoms)

Select all that apply:

☐ Full history
☐ Physical examination
☐ Assessment of mood
☐ Routine blood tests
☐ Infection-related blood tests
☐ Other blood testing
☐ Referral to another service
☐ Self-management advice without referral
☐ Social prescribing
☐ NHS Talking Therapies
☐ Watchful waiting/no active management
☐ Safety-netting advice
☐ Rehabilitation services
☐ Medication
☐ Other: _________

8. When considering whether symptoms may represent a PIC, which best describes your approach?

☐ I generally use a fixed timeframe as a guide
☐ I do not use a fixed timeframe but consider duration alongside other clinical features
☐ I have not considered this before
☐ Other: _________

If fixed timeframe:

☐ 4–8 weeks
☐ 8–12 weeks
☐ ≥12 weeks

If no fixed timeframe, factors influencing judgement:

☐ Duration of symptoms
☐ Severity/functional impact
☐ Symptom pattern
☐ Absence of alternative diagnosis
☐ Type of initial infection
☐ Patient concerns
☐ Other: _________

9. Coding practices

Which codes may you use for PIC consultations?

☐ I do not code these consultations
☐ Tired all the time
☐ Malaise
☐ Fatigue
☐ Chronic fatigue syndrome
☐ Chronic pain
☐ Post-viral fatigue
☐ Medically unexplained symptoms
☐ POTS
☐ Myalgic encephalomyelitis
☐ Post-infectious syndrome
☐ Post-COVID-19 syndrome
☐ Other: _________

10. Does your practice refer/signpost patients with PICs to community or external services?

☐ Yes
☐ No
☐ Not sure

If yes, services:

☐ Community rehabilitation services
☐ Community mental health services
☐ Health coaching/lifestyle support
☐ NHS self-referral or online self-help services
☐ Voluntary/third-sector services
☐ Other: _________

11. How confident do you feel managing patients with PICs?

☐ Very confident
☐ Fairly confident
☐ Not very confident
☐ Not at all confident

12a. To what extent do existing primary and community care services meet the needs of people living with PICs?

☐ Great extent
☐ Some extent
☐ Limited extent
☐ Not at all
☐ Not sure

12b. Please explain: _______________________

13a. To what extent could a personalised PIC management service delivered by a skilled primary care workforce meet patient needs?

☐ Great extent
☐ Some extent
☐ Limited extent
☐ Not at all
☐ Not sure

13b. Please explain: _______________________

14. Who would be best placed to deliver this service?

☐ GP
☐ Practice nurse/nurse practitioner
☐ Practice pharmacist
☐ Allied health professional
☐ Social prescriber/health and wellbeing coach
☐ Mental health practitioner
☐ Community rehabilitation team
☐ Other: _________
☐ Do not know

15. At what level would this service be best organised?

☐ Individual practice
☐ Primary Care Network/collaborative/federation
☐ Community services
☐ Integrated Care Board/System
☐ Hospital trust
☐ Not sure

15b. Please explain: _______________________

Survey 2: Other General Practice Healthcare Practitioner Survey

*(Practice nurses, advanced nurse practitioners, physiotherapists, pharmacists, physician associates, mental health practitioners, paramedics, allied health professionals etc.)*

Demographics

Practice name: ___________

Practice postcode: ___________

Role:
☐ Practice pharmacist
☐ First contact physiotherapist
☐ Practice nurse
☐ Advanced nurse practitioner
☐ Advanced clinical practitioner
☐ Practice mental health practitioner
☐ Physician associate/assistant
☐ Practice paramedic
☐ Other allied health professional
☐ Other

Number of patient-facing sessions per week
☐ <2 sessions
☐ 2-4 sessions
☐ 5-7 sessions
☐ >8 sessions

Main survey

Questions are equivalent to the GP survey but adapted for professional scope.

Key differences:

- Question 1: Number of patients seen with suspected PICs in previous month.
- Question 2: Frequency of PIC-associated symptoms.
- Question 4: Management approaches routinely suggested:
  - Standardised assessment tools
  - Individualised goal-setting
  - Pacing/energy management
  - Graded activity programmes
  - Psychological support
  - Self-management education
  - Referrals
  - Communication with GP team
  - Peer/community support
- Question 5: Recording/coding practices.
- Question 6: Referral/signposting pathways.
- Question 7: Confidence managing PICs.
- Question 8: Whether current services meet patient needs.
- Question 9: Views on personalised PIC management service.
- Question 10: Who should deliver service.
- Question 11: Best organisational level.

Exit questions (both surveys)

Would you be interested in your practice being involved as a trial site if funding was secured?

☐ Yes
☐ Maybe / would like more information
☐ No
☐ Not my role to agree to research

Contact details (optional): ___________

Would you like your name acknowledged in publications/reports?

☐ Yes
☐ No

If yes, name for acknowledgement: ___________

Prize draw (£100 shopping voucher):

☐ Yes
☐ No

Email (if yes): ___________

**Supplementary Table S3. Data extraction table.**

| **Author & Year** | **URL** | **Study Type** | **PIC(s)** | **Type of Primary Care Approach Studied** | **Approach Description** | **Role of Primary Care in the Study** | **Self-Management Support Provided** | **Patient Self-Management Reported** | **Setting / Context** | **Sample Size** | **Sample Characteristics** | **Key Findings** | **Equity Findings** | **Barriers to Care** | **Enablers to Care** |
| --- | --- | --- | --- | --- | --- | --- | --- | --- | --- | --- | --- | --- | --- | --- | --- |
| **NHS England (2021)** | <https://www.england.nhs.uk/coronavirus/wp-content/uploads/sites/52/2021/06/C1312-long-covid-plan-june-2021.pdf> | Policy report | Long COVID (post–COVID-19) | National service model | National primary care-led pathway; referral to LC clinics; integrated MDT care | Primary care as first contact, assessor, coordinator, referrer | Digital recovery programme (Your COVID Recovery); clinician-supported self-management; education resources | N/A | UK primary care, PCNs, community, ICS pathways | N/A | N/A | Establishes structured pathways; emphasises GP coordination and self-management | Recognises inequalities (ethnicity, deprivation); limited subgroup detail | Service variation; access issues; GP workload; digital exclusion | National funding; training; integrated pathways; digital tools |
| **Alwan et al. (2023)** | <https://journals.plos.org/plosone/article?id=10.1371/journal.pone.0284297> | Mixed-methods protocol | Long COVID (post–COVID-19) | Community outreach intervention (planned) | Identify undiagnosed individuals; link to GP, social prescribing, services | Primary care as referral endpoint and care access point | Planned personalised support via social prescribing and education | N/A | Community-based outreach linked to primary care | n≈20-30 (planned) | Adults ≥18; underserved groups targeted (planned) | N/A | N/A | Anticipated: stigma, access barriers, dismissal | Anticipated: community partnerships; trusted organisations; tailored outreach |
| **Sivan et al. (2022)** | 10.1136/ bmjopen-2022-063505 | Mixed-methods protocol | Long COVID (post–COVID-19) | Integrated care pathway optimisation (planned) | QI collaborative; digital monitoring; pathway redesign; MDT care | Primary care involved in pathway redesign and data (GP cohort) | Planned digital monitoring; self-management tools; co-designed support | N/A | Primary care + LC clinics + home monitoring | n≈5100 (planned) | Adults ≥18; clinic and GP populations (planned) | N/A | N/A | Anticipated: variable access; long waits; GP uncertainty | Anticipated: patient co-design; tech-enabled monitoring; QI cycles |
| **Brennan et al. (2022)** | [10.3399/BJGPO.2021.0178](https://doi.org/10.3399/BJGPO.2021.0178) | Scoping review | Long COVID (post–COVID-19) | Synthesis of primary care approaches | Review of GP-led assessment, monitoring, referral, holistic care | GP central in assessment, coordination, referral | Education, reassurance, symptom monitoring, community support | N/A | UK primary care | N/A | Adults ≥18 | Highlights need for holistic, continuous GP care and MDT access | N/A | GP uncertainty; lack of guidance; service variation | Training; standardised tools; MDT collaboration |
| **Bulley et al. (2022)** | [10.1136/bmjopen-2021-056568](https://doi.org/10.1136/bmjopen-2021-056568) | Mixed-methods study | Long COVID (post–COVID-19) | Patient experience informing service recommendations | Co-produced recommendations for multidisciplinary, accessible support | Primary care as access point and coordinator | Education; peer support; rehabilitation advice; online/telephone support | Engagement with peer support; seeking info; coping strategies | Community and primary care-linked | n=675 survey; n=107+ qualitative subsamples | Adults ≥18; majority female; limited diversity | Identifies unmet needs; need for personalised, accessible support | Digital exclusion; low-income barriers | Limited services; funding; digital exclusion; access inequality | Peer support; flexible delivery; stakeholder involvement |
| **Busse et al. (2025)** | 10.1136/bmjmed-2024-001068 | RCT | Long COVID (post–COVID-19) | Intervention (self-management programme) | Personalised one-to-one sessions; goal-setting; self-efficacy support | Complements primary care; recruited via GP/community | Structured self-management programme; handbook; coaching | Goal-setting; problem-solving; symptom management | Community/outpatient | n=554 (277 intervention; 277 control) | Adults ≥18; mean age 50; 72% female; 92% White | Improved fatigue, QoL, self-efficacy; no primary outcome difference | Digital exclusion | Variation in usual care; digital barriers | Personalisation; flexible delivery |
| **Combet et al. (2025)** | [10.1038/s41591-024-03384-x](https://doi.org/10.1038/s41591-024-03384-x) | RCT | Long COVID (post–COVID-19) | Intervention (weight management) | Remote diet programme; professional support; symptom-targeted | Indirect (usual care comparator; GP-linked recruitment possible) | Structured diet; behavioural support; remote guidance | Adherence to diet; engagement with programme | Remote/home-based | n=234 (116 intervention; 118 control) | Adults ≥18; mean age ~46; 85% female; 91% White | Improved symptoms, QoL, BP; effective weight loss | Lower completion in deprived groups | Attrition in deprived groups | Flexible delivery; structured support |
| **Cooper et al. (2024)** | [10.1136/bmjopen-2023-082830](https://doi.org/10.1136/bmjopen-2023-082830) | Qualitative | Long COVID (post–COVID-19) | Patient & GP experience of care | Explores perceptions of community rehabilitation | Primary care as referral and coordination point | N/A | Self-advocacy; seeking care; managing symptoms | Community rehab + GP | n=24 (11 PwLC; 13 GPs) | Adults ≥18; PwLC mean age ~53; 91% female PwLC; ethnicity not reported | Highlights need for coordination and person-centred care | N/A | Unclear pathways; limited knowledge; access issues | Positive rehab experiences; integrated care desire |
| **Darbyshire et al. (2024)** | 10.1016/j.clinme.2024.100237 | Mixed-methods QI | Long COVID (post–COVID-19) | Service model improvement | Tiered MDT model; QI collaborative; standardised guidance | Primary care in tiered model; triage and management | Guidance; pacing; PROMs; education; vocational support | Symptom tracking; pacing; engagement with care | Primary + secondary integrated clinics | n=59 (29 patients; 30 HCPs) | Adults ≥18; mixed patient/HCP sample; ethnicity variably reported across sites | Improved coordination; shared learning; holistic care | Improved access for some underserved groups | Complex pathways; funding; digital barriers | QI collaboration; MDT care; patient involvement |
| **Fang et al. (2024)** | [10.1186/s12913-024-10891-7](https://doi.org/10.1186/s12913-024-10891-7) | Qualitative longitudinal | Long COVID (post–COVID-19) | Patient experience of care pathways | Explores fragmented care and evolving services | Primary care as gateway; limited follow-up | Some psychology-led input | Self-monitoring; proactive follow-up; advocacy | UK primary + specialist pathways | Phase 1 n≈92; Phase 2 n≈86 | Adults 18-79; ~72% female; ~55% White British; ~37% South Asian | Fragmented care; limited access; evolving holistic services | Ethnic diversity; limited Black inclusion | Poor access; funding instability; navigation difficulty | Patient-centred care; proactive engagement |
| **Forshaw et al. (2023)** | [10.1371/journal.pone.0272472](https://doi.org/10.1371/journal.pone.0272472) | RCT protocol | Long COVID (post–COVID-19) | Intervention (integrated care pathway) | Cluster RCT testing integrated pathway vs usual care; digital rehab + imaging | Primary care networks recruit and manage pathway allocation | Digital rehab (Living With COVID Recovery); structured care pathways | N/A | Primary care networks + LC clinics | n≈1130 (planned) | Adults ≥18; LC ≥12 weeks (planned) | N/A | N/A | Anticipated: digital barriers | Anticipated: integrated pathways; structured system design |
| **Fowler-Davis et al. (2021)** | [10.3390/ijerph182413191](https://doi.org/10.3390/ijerph182413191) | Mixed-methods | Long COVID (post–COVID-19) | Intervention (community MDT model) | Co-produced virtual MDT rehabilitation (3 sessions) | GP informed and linked to MDT intervention | Pacing; goal-setting; symptom monitoring; wellbeing tracking | Self-management engagement during programme | Community MDT linked to primary care | n=10 | Adults 38-75; 80% female; 40% White; 60% Asian or Black British | Improved QoL, fatigue; high acceptability | Deprived community targeted; improved access but gaps remain | Digital barriers; low engagement history | Co-production; trusted community links |
| **Welsh Gov (2022)** | https://cedar.nhs.wales/files/adferiad-recovery-long-covid-service-national-evaluation-v1-3-pdf/ | Service evaluation | Long COVID (post–COVID-19) | National service evaluation | Evaluation of national LC rehabilitation hubs | GP referral pathway to LC hubs | Rehabilitation advice; peer support; group education | Engagement with rehab sessions; coping strategies | NHS Wales LC hubs + primary care | n=589 | Adults ≥18; 66% female; 96% White British | Improved QoL in discharged group; high satisfaction | N/A | Delayed referral; GP awareness gaps | Empathetic care; group support; structured service |
| **Greenhalgh et al. (2024)** | [10.1111/1467-9566.13819](https://doi.org/10.1111/1467-9566.13819) | Ethnography | Long COVID (post–COVID-19) | Service model (clinical practice) | MDT clinic decision-making and relational care | Primary care refers; clinics interpret and manage uncertainty | Pacing; breathing exercises; mindfulness; online resources | Patient engagement with pacing and self-monitoring | UK LC clinics (hospital/community interface) | n=294 patients; n=45 MDT meetings | Adults 18-87; median 48; 61% female; ethnicity not fully reported | Care shaped by uncertainty; relational knowledge key | N/A | Variable access; uncertainty; knowledge gaps | MDT deliberation; patient-centred reasoning |
| **Greenhalgh et al. (2020)** | <https://doi.org/10.1136/bmj.m3026> | Narrative review | Long COVID (post–COVID-19) | Primary care guidance model | GP-led holistic symptom management framework | GP central in monitoring and referral | Pacing; self-monitoring; breathing exercises; education | Patient self-monitoring and graded activity | Primary care/community | N/A | N/A | Early recognition of LC; role of holistic GP care | N/A | Uncertainty in diagnosis; lack of testing | Telehealth; holistic care principles |
| **Haag et al. (2023)** | [10.3310/nihropenres.13315.2](https://doi.org/10.3310/nihropenres.13315.2) | RCT protocol | Long COVID (post–COVID-19) | Intervention (weight management) | Remote diet programme; professional support; symptom-targeted | GP/community recruitment pathways | Structured diet programme; behavioural support | Engagement with dietary programme | Remote/home-based | n≈240 (planned) | Adults ≥18; BMI ≥27 (≥25 South Asian) (planned) | N/A | N/A | Anticipated: adherence challenges | Anticipated: remote delivery; tailored support |
| **Harenwall et al. (2021)** | [10.1177/21501319211067674](https://doi.org/10.1177/21501319211067674) | Quantitative | Long COVID (post–COVID-19) | Intervention (rehabilitation programme) | 7-week psychology-led virtual rehab | GP signposting into rehab pathway | CBT-informed rehab; pacing; activity management | Self-monitoring; pacing; behaviour change strategies | Primary care wellbeing service | n=149 baseline; n=76 follow-up | Adults ≥18; mean age ~47-49; ~72% female; ~71% White British | Improved HRQoL and self-management confidence | Some BAME differences in baseline severity | Digital barriers; attrition | Structured MDT rehab; virtual delivery |
| **Harenwall et al. (2022)** | [10.3390/jcm11206214](https://doi.org/10.3390/jcm11206214) | Quantitative | Long COVID (post–COVID-19) | Intervention (cohort analysis) | 7-week psychology-led virtual rehab programme examining psychological mechanisms | Embedded in primary care rehab service | Psychological support targeting PTSS | Engagement in therapy-based self-management | Virtual rehab service | n=154 baseline; n=79 follow-up | Adults ≥18; mean age ~47; ~87% female; ~73% White British | PTSD + breathlessness interaction worsens fatigue | N/A | PTSS barriers impact engagement | MDT psychological input |
| **Kalfas et al. (2024)** | [10.1111/hex.14108](https://doi.org/10.1111/hex.14108) | Qualitative | Long COVID (post–COVID-19) | Patient experience of care | Exploration of care gaps and lived experience | GP as first access point | N/A | High self-management reliance; online resources use | Primary care access | n=19 | Adults ≥18; mean age ~50; 68% female; ~90% White | Dismissal and lack of support common; identity impact | N/A | Symptom dismissal; poor GP access | Peer support; acceptance strategies |
| **Mansoubi et al. (2025)** | [10.1136/bmjopen-2024-094658](https://doi.org/10.1136/bmjopen-2024-094658) | Survey | Long COVID (post–COVID-19) + ME/CFS (unspecified) | Patient experience of care | Comparison of healthcare experiences across conditions | GP key entry point; limited ongoing support | N/A | Strong self-management reliance (pacing, rest) | Community | n=10458 | Adults ≥18; 83% female; 95% White | Poor satisfaction; long diagnostic delays; high burden | N/A | Delayed diagnosis; poor NHS support | Online peer support; self-management reliance |
| **Mir et al. (2025)** | [10.1111/hex.70336](https://doi.org/10.1111/hex.70336) | Mixed-methods | Long COVID (post–COVID-19) | Service experience + access pathways | Exploration of inclusive service development and barriers | GP referral | Peer support; information provision; advocacy support | Self-advocacy; navigating fragmented systems | GP practices + LC clinics + community | n=41 (23 PwLC; 18 experts) | Adults ≥18; 74% female; 35% White; 30% South Asian; 17% Black | Fragmented care; under-referral; access inequalities | Strong inequities by ethnicity and SES | Poor access; mistrust; fragmented pathways | Integrated care; third-sector collaboration |
| **Mullard et al. (2024)** | 10.1111/1467-9566.13795 | Qualitative | Long COVID (post–COVID-19) | Patient experience of diagnostic pathway | Exploration of “diagnostic odyssey” and stigma | GP gatekeeping | Informal self-management; peer/family support | High self-advocacy and informal coping | Primary care | n=41 (23 PwLC; 18 experts) | Adults ≥18; 74% female; 35% White; 30% South Asian; 17% Black | Diagnostic delay; invalidation; emotional burden | Strong inequities by ethnicity and SES | GP dismissal; lack of pathways; stigma | Peer support; self-advocacy |
| **Morrow et al. (2022)** | [10.1186/s13063-022-06632-y](https://doi.org/10.1186/s13063-022-06632-y) | RCT protocol | Long COVID (post–COVID-19) | Intervention (exercise rehabilitation) | 12-week resistance exercise programme | GP recruitment and follow-up coordination | Exercise guidance; remote support calls | Home-based exercise adherence | Primary care + community | n≈220 (planned) | Adults ≥18; post-acute COVID (≤6 months) | N/A | N/A | Anticipated: adherence challenges | Anticipated: simple home-based design; regular contact |
| **Nurek et al. (2021)** | 10.3399/BJGP.2021.0265 | Delphi | Long COVID (post–COVID-19) | Primary care management guidance | Consensus recommendations for GP-led assessment and care | GP central in diagnosis, monitoring, referral | Pacing; education; symptom monitoring; social prescribing | Patient pacing and symptom self-monitoring | Primary care + MDT networks | n=33 | Clinicians (UK multidisciplinary) | Consensus on holistic GP-led management and MDT care | N/A | Limited evidence base; service variation | GP coordination; MDT collaboration |
| **Parkin et al. (2021)** | [10.1177/21501327211010994](https://doi.org/10.1177/21501327211010994) | Service description | Long COVID (post–COVID-19) | Service model (3-tier pathway) | Tiered MDT rehabilitation system integrated with GP | GP referral and triage into tiers | Rehabilitation advice; online resources; pacing | Engagement with tiered rehab pathway | Primary + community care | n=225 | Adults ≥18; mean age 48; 68% female; 74% non-hospitalised | Feasible MDT pathway; high demand; complex cases common | N/A | Capacity constraints; variation in referrals | Strong GP linkage; MDT coordination |
| **Razai et al. (2021)** | [10.1177/21501327211041846](https://doi.org/10.1177/21501327211041846) | QI survey | Long COVID (post–COVID-19) | Patient experience of GP care | QI survey of GP experiences and access barriers | GP as first contact; often reactive care | N/A | Self-management due to limited support | GP practices | n=41 | Adults 19-82; 66% female; 56% White; 22% Asian; 15% Black | Poor access; lack of follow-up; isolation | Digital exclusion and access inequality | Poor GP access; lack of continuity | Desire for proactive GP care |
| **Skilbeck et al. (2023)** | [10.1186/s41687-023-00570-2](https://doi.org/10.1186/s41687-023-00570-2) | Qualitative | Long COVID (post–COVID-19) | Integrated psychological service model | Experience of NHS primary care psychology support | GP referral into psychology service | CBT; coping strategies; symptom tracking | High self-management reliance | Primary care mental health service | n=18 | Adults ≥18; 72% female; 67% White; 17% Asian; 11% Black | Identity disruption; uncertainty; gradual adaptation | N/A | Feeling dismissed; uncertainty | Psychological support; acceptance strategies |
| **Smith et al. (2023)** | [10.3389/fmed.2023.1149922](https://doi.org/10.3389/fmed.2023.1149922) | Quantitative | Long COVID (post–COVID-19) | Intervention (blended rehab) | 12-week digital + community rehabilitation programme | Indirect GP referral/self-referral pathways | Exercise prescription; coaching; digital platform | Structured exercise adherence; monitoring | Community fitness + digital platform | n=601 | Adults ≥18; mean age 47; 77% female; 89% White British | Improved HRQoL, function, reduced healthcare use | N/A | Digital literacy barriers; access inequity | Blended delivery; coaching support |
| **Sunkersing et al. (2024)** | [10.1136/bmjopen-2023-080967](https://doi.org/10.1136/bmjopen-2023-080967) | Qualitative | Long COVID (post–COVID-19) | Service experience | Experiences of LC clinic care and GP access | GP referral into LC clinics; variable access | Peer support; education; coping strategies | Engagement with peer groups and coping tools | Primary care + LC clinics | n=36 (21 PwLC; 15 HCPs) | PwLC: 86% female; 57% aged 38-47; 57% White British; 14% Asian | LC clinics valued but access inconsistent | Ethnic diversity present but unequal access persists | Long waits; staffing shortages; GP dismissal | MDT clinics; peer support |
| **Vanova et al. (2024)** | [10.1186/s13063-024-08554-3](https://doi.org/10.1186/s13063-024-08554-3) | RCT protocol | Long COVID (post–COVID-19) | Intervention (cognitive rehabilitation) | Telehealth cognitive rehab (10 sessions) | GP/community referral into memory clinic pathway | Cognitive strategies; goal-setting; pacing | Home-based cognitive exercises | NHS memory clinics | n≈120 (planned) | Adults ≥18; working-age; cognitive impairment (planned) | N/A | N/A | Anticipated: digital exclusion; fatigue burden | Anticipated: remote flexible delivery |
| **Walker et al. (2023)** | [10.1136/bmjopen-2022-069217](https://doi.org/10.1136/bmjopen-2022-069217) | Observational | Long COVID (post–COVID-19) | Clinical outcomes study | Observational study of LC clinic patients | GP referral into LC clinics | Symptom monitoring via PROMs | Engagement with digital symptom reporting | LC clinics | n=3754 | Adults ≥18; mean age 48; 71% female; 87% White | Fatigue strongest predictor of impairment | N/A | Digital selection bias | PROM-based monitoring system |
| **Williams et al. (2024)** | 10.1101/2024.10.25.24316101 | Mixed-methods | Long COVID (post–COVID-19) | Service implementation evaluation | GP-led MDT community LC service | GP triage and coordination | Goal-setting; education; symptom monitoring via PROMs | Engagement with digital symptom reporting | Community LC service | n=116 + n=11 interviews | Adults ≥18; mean age 51; 71% female; 77% White British | Improved reassurance; high onward referral needs | N/A | Funding and capacity constraints | MDT teamwork; integrated pathways |

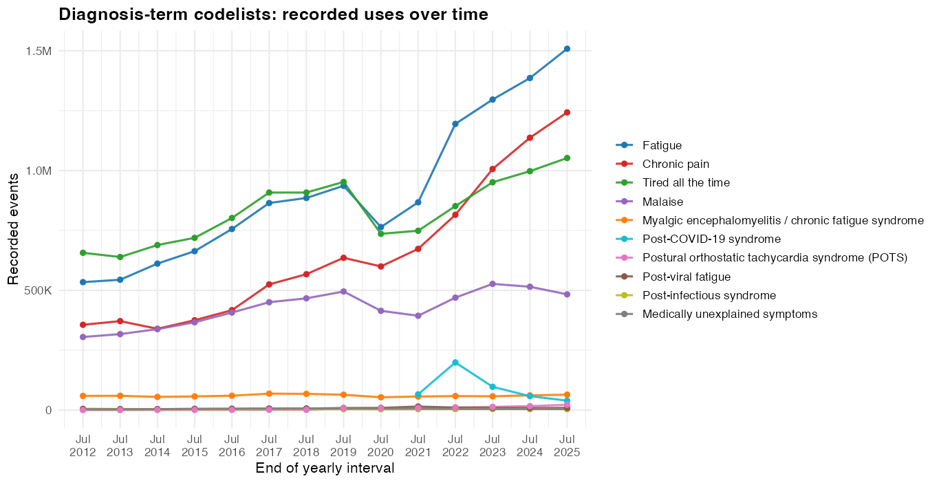
**Supplementary Figure S4. Annual recorded use of SNOMED CT codes corresponding to post-infection condition (PIC)-related diagnostic terms in English primary care, 2011-2025.**

Annual counts were derived from NHS England primary care SNOMED-CT code usage data. Codelists were developed from diagnostic terms reported by survey respondents as being used to code PICs and comprised 252 SNOMED-CT concepts across 10 diagnostic categories. Each line represents the total annual recorded use of all codes within a codelist, irrespective of clinical indication; counts are therefore not specific to PICs. Reporting intervals end in July. Counts below five were suppressed by the data source, and all other counts were rounded to the nearest ten. In 2024/25, these codelists accounted for approximately 4.5 million recorded code uses in English primary care. The most frequently recorded categories were fatigue (approximately 1.5 million uses), chronic pain (1.2 million), "tired all the time" (1.1 million), and malaise (483,270). Coding for these symptom-based categories generally increased between 2011 and 2025, with a temporary reduction between 2020 and 2021. In contrast, diagnosis-specific terms were recorded less frequently; myalgic encephalomyelitis/chronic fatigue syndrome was recorded 64,620 times in 2024/25, while post-Covid-19 syndrome coding peaked at nearly 200,000 uses in 2021/22 before declining to 39,640 in 2024/25.
